# Digitisation and linkage of PDF formatted 12-lead ECGs in Adult Congenital Heart Disease

**DOI:** 10.1101/2024.12.16.24319092

**Authors:** Muhammet Alkan, Fani Deligianni, Christos Anagnostopoulos, Idris Zakariyya, Gruschen Veldtman

## Abstract

**BACKGROUND:** 12-lead ECG’s form an essential part of the late follow-up of adults with congenital heart disease (ACHD). Such ECGs are most frequently reviewed by clinicians in paper or PDF formats. These visual representations of the original vector data do not easily lend themselves to be directly analysed with the increasingly powerful Machine Learning algorithms that hold promise in risk prediction and early prevention of adverse events.

**OBJECTIVES:** In this work, we set out to recreate the original digital signals from ECG PDF documents by a series of data processing steps, validate accuracy of the process, and demonstrate its potential utility in research.

**METHODS:** Using 4153 ECG PDF documents from 436 ACHD patients, we created a “pipeline” to successfully digitise the visually represented ECG vector datasets. We then proceed with the validation of the digitised ECG dataset using several features that are also calculated by the vendor, such as QRS duration, PR interval and ventricular rate, on all the patients.

**RESULTS:** We confirmed a strong correlation with the vendor measured ECG parameters including PR interval (*R* = 0.941, *P* < 0.05), QRS duration (*R* = 0.949, *P* < 0.05) and ventricular rate (*R* = 0.971, *P* < 0.05). Further, using Support Vector Machine (SVM), a well-established Machine Learning (ML) model we demonstrate the ability of the digitised ECG dataset to accurately predict anatomic diagnosis in ACHD.

**CONCLUSIONS:** Digitisation of PDF formatted ECG signal data can be accomplished with good accuracy and can be used in clinical research in ACHD.

## Introduction

With advances in medical sciences, more than 90% of patients with even the most complex congenital heart disease (CHD) survive into adult life^1^. With this transformational success of medical and surgical advances, there has emerged a growing burden of morbidity and mortality as individuals enter later into adult life. Increasing attention is being drawn to the ability to predict which individuals are likely to deteriorate, and those at risk of mortality^2,3,4,5,6^. Electrocardiograms (ECGs) are a fundamental aspect of such assessments^7,8,9,10^. They have been demonstrated to carry important diagnostic and prognostic information that correlate with clinically important outcomes^10,11,12,13,14^. For example, in CHD, clinicians use derived ECG measures such as QRS duration, QRS fragmentation, QRS axis and R-wave height along with PR intervals as indicators of underlying anatomy and prior surgery, disease state and for mortality prognosis in some specific settings^29^.

More recently Machine Learning (ML) algorithms promise to extract relevant ECG features and exceed the performance of more traditional and more manual approaches in disease diagnosis and prognostication^15,16^. In a recent review, Helman et al.^17^ highlighted that the development of ML algorithms to process 12- lead ECGs for CHD is a promising direction, which currently remains unexplored due to the lack of diverse and large datasets.

In our previous work, we have identified the opportunity to develop digital twins of ACHD patients by extracting information over a decade from clinical letters, which involved diagnoses, diagnostic complexity, interventions, arrhythmia, medications, and demographic data^18^. We believe this information can also be linked to the patient’s 12-lead ECG dataset when in digitised format. This provides the opportunity to develop robust ML models for diagnoses and risk prediction and explore their capabilities.

In clinical practice, 12-lead ECGs are often interpreted from scanned in or printed PDF documents. Valuable information can be derived from such visual manual assessment, such as cardiac rhythm and rate, haemodynamic characteristics like pulmonary hypertension, and prognostic information relating to sudden cardiac death risk. Machine learning on the actual raw signal data enhance further diagnostic and prognostic capacity. Due to proprietary software and analysis of raw ECG signal data, such “raw” data is not readily available for de-novo or empiric data analysis or use in further research. Being able to accurately reproduce the digitised ECG signals from PDF documents can potentially enable signal data to be used in developing ML models for risk stratification and diagnosis.

For ECG digitisation, all current approaches focus on image processing (i.e. on scanned images or PDF documents converted into a desired image format) rather than the conversion to the vector data represented by the PDF document^19,20^, and this in particular has not previously been evaluated in CHD.

We propose a stepwise approach as a pipeline that assemble several steps that are combined to extract 12-lead ECG digital signals from ECG PDF documents. We adopt open-source generic Python libraries to extract information about the text and the geometric objects present in ECG PDF documents, e.g., PyMuPDF^21^. We then validate the digitised dataset using derived ECG features such as QRS duration, PR interval and heart rate, and further demonstrate its potential research utility by using it to predict anatomic diagnosis associated with CHD. This is an initial step towards mortality prediction, which requires significantly larger numbers of subjects to achieve.

## Methods

### Linkage of CHD EHR

Study approval was obtained by the Institutional Governance Division of the NHS Golden Jubilee National Hospital. Demographic information including age, gender, anatomic diagnoses, and prior surgical intervention were extracted from clinical letters as previously described in^18^. The most common condition was Tetralogy of Fallot, in 197 patients, followed by Pulmonary Atresia, in 96 patients. The top 15 anatomic diagnoses are summarised in Figure 1.

**Figure 1:**
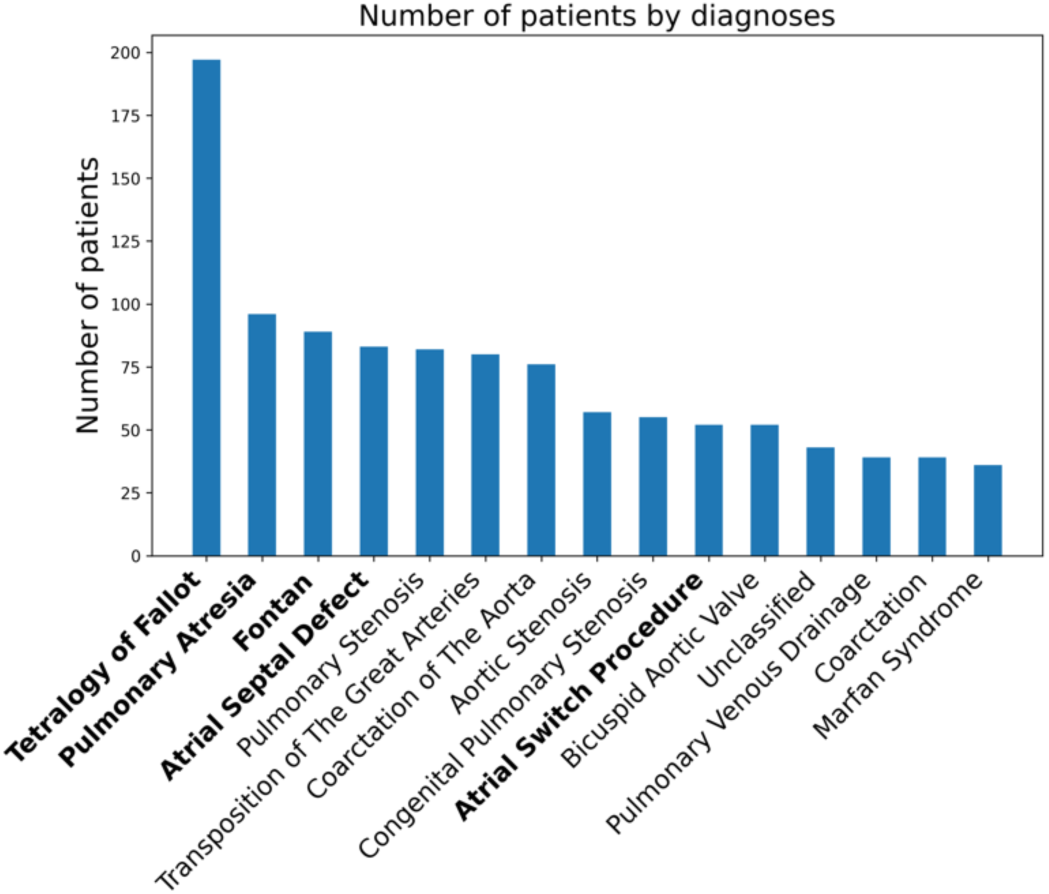
Top 15 anatomic CHD diagnoses for 1409 patients. Summary of all the patients initially screened for inclusion, only the top 15 anatomic CHD diagnoses are shown.

Among these conditions, we have selected the 5 most common conditions (shown in bold in Figure 1) for diagnosis prediction including:

1. Tetralogy of Fallot (ToF) (Diagnosis List)
2. Pulmonary Atresia (PA) (Diagnosis List)
3. Fontan (Intervention & Diagnosis List)
4. Atrial Septal Defect (ASD) (Diagnosis List)
5. Mustard (Intervention List) including other atrial switch procedures.

Information from clinical letters was linked to ECG PDFs based on the patient chart number. Patients were excluded if they did not have available ECG’s or were in a permanent dysrhythmia, such as atrial fibrillation or flutter, or were atrioventricular paced. This led to 436 patients being included in our investigation. Of these 436 patients, 173 patients had ToF, 77 patients had ASD, 73 patients had PA, 66 patients had Fontan and 47 patients had Mustard. Overall, 4153 ECG PDF documents were extracted from these 436 patients with diagnoses as outlined.

### ECG Data Preprocessing

12-lead ECG data were extracted from ECG PDF documents obtained via the Marquette™ 12SL by GE Healthcare analysis program^22^. For resting ECGs, the analog voltage potential is digitised into 4.88-μV units at a rate of 4kHZ. The software down-samples the signal to 500 samples per second and represents a value every 0.05mm on a chart. The ECG signal has been pre-processed to remove noise and QRS template matching was employed to extract ECG features and export ECG waveforms to PDF documents in a vectorised format. Each PDF document contains 12 leads in a specific order (Lead I, II, III, aVF, aVR, aVL, V1, V2, V3, V4, V5, V6), and provides only 2.5-second strip for each lead.

### ECG Digitisation Process

We developed a new algorithm using vector drawings on the ECG PDF documents to digitise ECGs without user intervention. PyMuPDF^21^ library was used to extract information about the graphics points present in all the PDF documents. The algorithm follows the steps detailed in Algorithm 1. An example ECG document with two of the extracted leads is shown in Figure 2.

**Figure 2:**
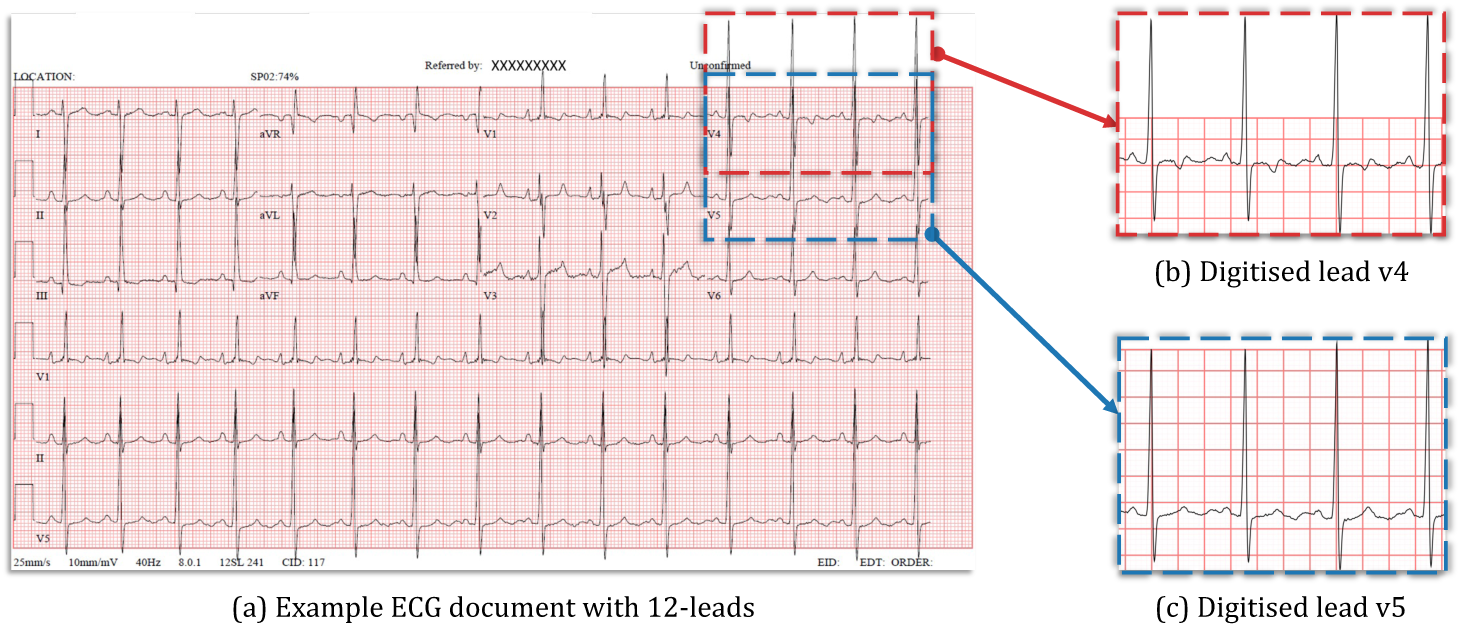
Digitisation and linkage of ECG PDF documents. On the left panel, a sample ECG PDF document that uses vectorised graphic format to store ECG waveforms are displayed. On the right panel, the results of the digitised leads (for lead v4 and v5) are displayed.

### Digitised ECG Analysis & ECG Peak Alignment

To facilitate analysis of ECG data with ML algorithms, we standardised them by *aligning* the ECG’s so that the peaks of the R-waves intersected and QRS complexes were digitally synchronised across all leads. An example of the average signal of the aligned ECGs for Mustard is shown in Figure 3 and its characteristic of the underline abnormality.

**Alg.1:**
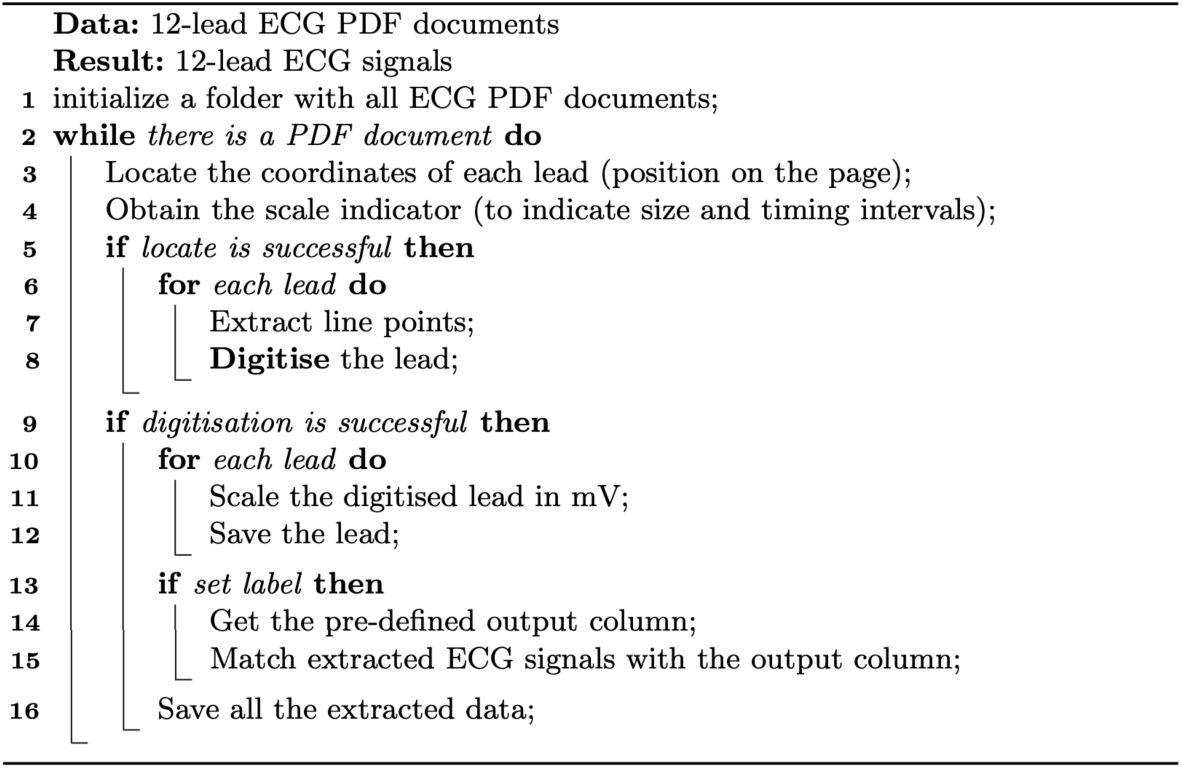
**ECG Digitisation Pseudocode** that shows each algorithmic step.

**Figure 3:**
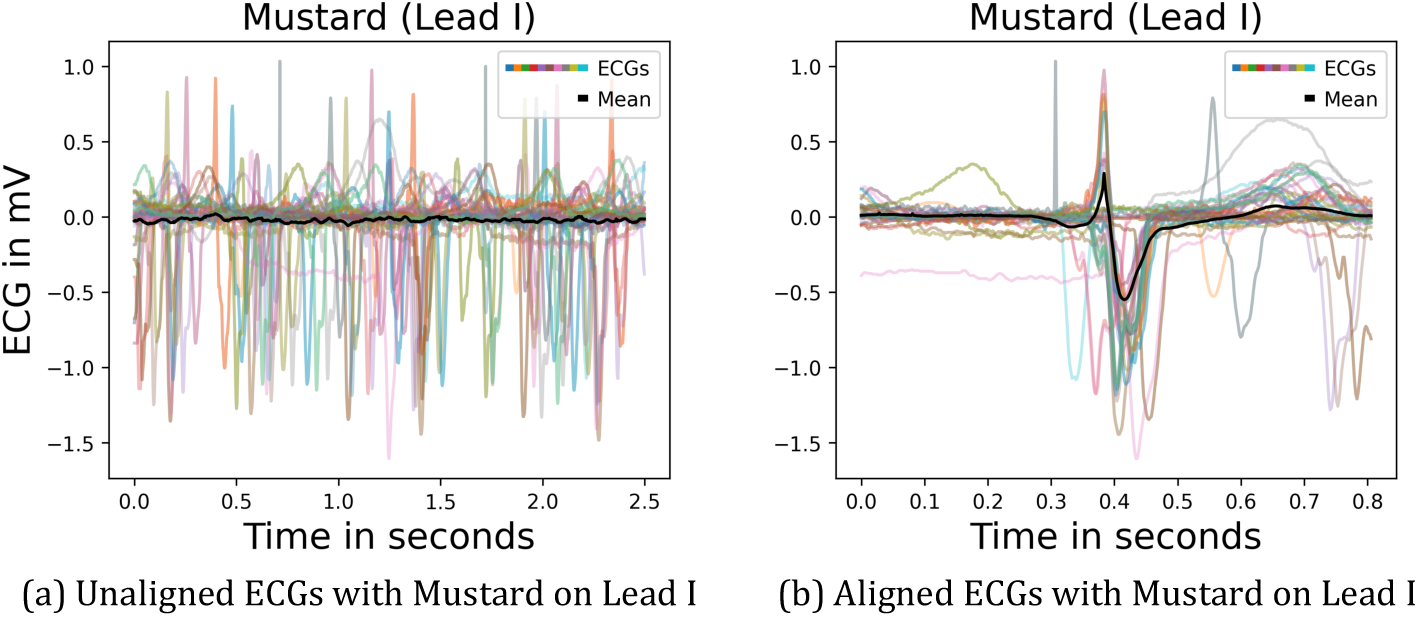
ECG alignment using a random 50 ECGs. ECG alignment using a random 50 ECGs on lead I of patients with a Mustard procedure for Transposition of the great arteries. On the left panel, individual non-aligned ECGs for the random 50 patients are displayed. On the right panel, QRS aligned ECGs with the overall signal vector (outlined in bold) for the same random 50 patients are displayed.

For validation of our Algorithm 1, we compared vendor derived ECG intervals with derived intervals from the digitally aligned extracted ECG signals. All the patients were checked to assess the accuracy of the digitisation algorithm.

The onsets and offsets for the points P, Q, R, S, and T in all 12 leads should be determined first to able to calculate the metrics like QRS duration, PR interval and ventricular rate. An example illustration can be found in Figure 4, on a single beat. In the vendor algorithm^22^, onsets are defined as the earliest deflection in any 12 leads, and offsets are defined as the latest deflection in any 12 leads. The QRS duration is measured in milliseconds from the earliest Q onset in any lead to the latest S offset in any lead. Similarly, the PR interval is measured in milliseconds from the earliest P onset in any lead to the QRS onset (or earliest Q onset) in any lead. For the ventricular rate (beats per minute), the number of beats is counted and divided by the time difference in minutes between the first and last beat. For all our corresponding calculations of QRS duration, PR interval and ventricular rate, the NeuroKit2^23^ library was used. We followed a processing pipeline similar to the vendors guidelines^22^ to enable meaningful comparison of the final measurements.

**Figure 4:**
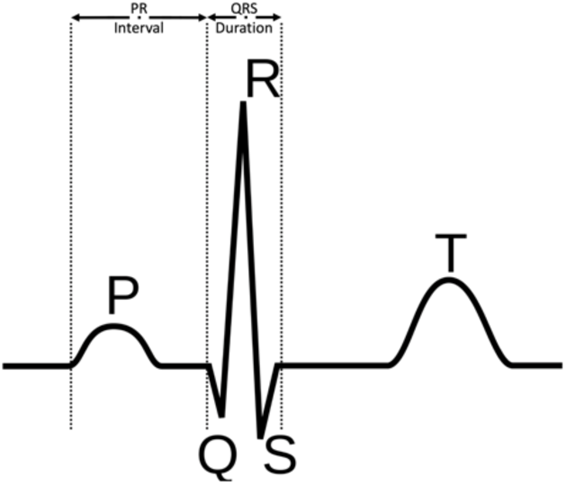
P, Q, R, S, and T points on a single ECG wave. Demonstration of a normal single lead ECG trace, with P, Q, R, S, and T points.

### Training to Predict Diagnosis

The Support Vector Machine (SVM) is a well-established ML model. The training of SVM model was performed on the ECG aligned time-series data to predict the corresponding diagnoses. SVM with the radial basis function (RBF) kernel, also known as the Gaussian kernel, in the scikit-learn package was used. Regularization is applied using the parameter C=1 with no limit on iterations, which permits a certain degree of tolerance for misclassification. The SVM model works by mapping training data into a high dimensional space to maximise the gap between the points of different labels^24^. When new data are received, they are mapped into the same space and then the SVM model makes a prediction based on a distance metric.

The training-testing split of data was repeated 100 times based on pseudo-randomised, stratified patient leave-out evaluation to ensure that the testing set of patients was representative for all the classes. One patient from each class was randomly selected to populate the testing set, and all the remaining patients to populate the training set. For each run, SVM model trained on the populated training set and then tested on the corresponding testing set that does not include any data from the patients in the training set (i.e., stratified patient leave-out method). Using such an approach, summarised in Figure 5, stratified patient leave-out enhances the capability of the SVM model to predict unseen data by reducing bias^25^. This approach ensures that each class is proportionally represented in both the training and test sets, thereby preventing overfitting and underfitting.

**Figure 5:**
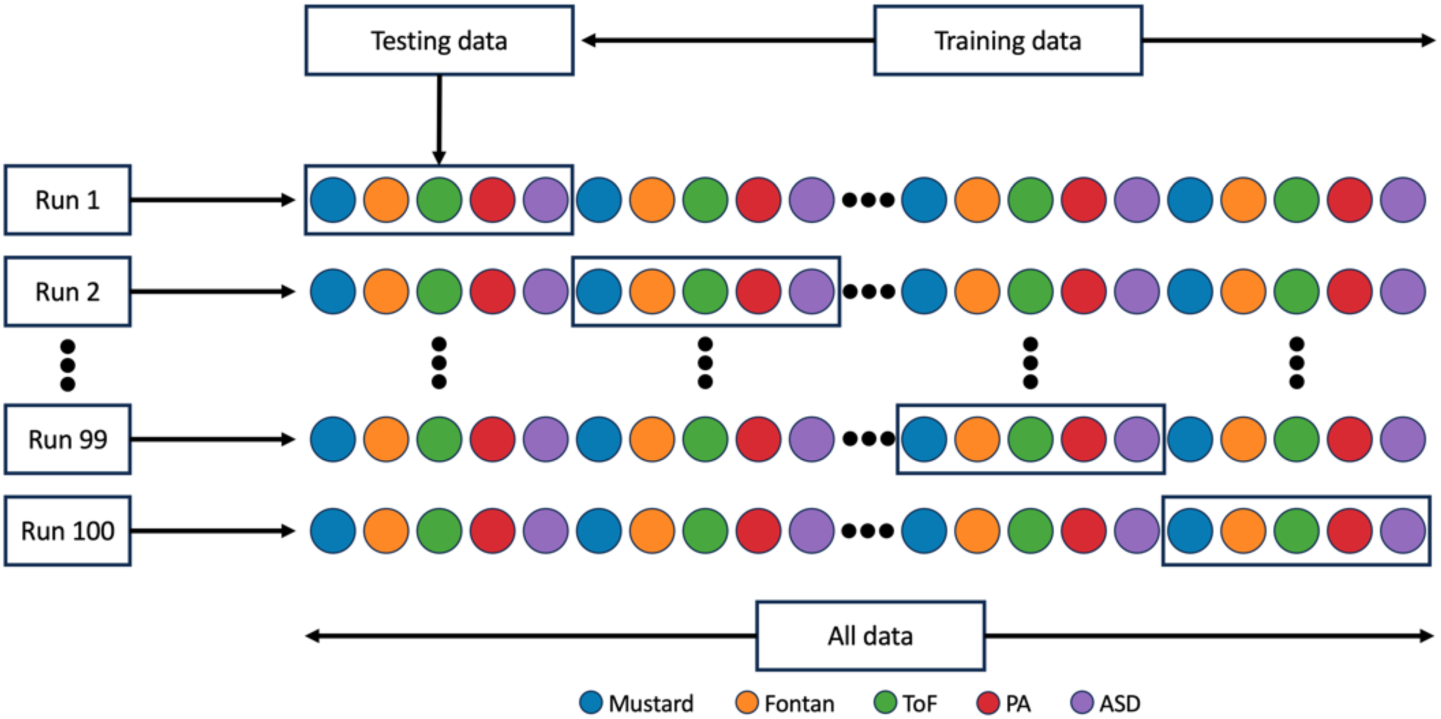
Stratified patient leave-out setup. Each colour represents a different diagnosis. Each coloured circle contains all the ECGs of a single patient with the corresponding diagnosis highlighted with the colour. Each run represents a training and testing cycle and the SVM model trained from scratch in each run. For each run, 5 patients randomly selected and all the data for these patients are kept for testing, while the rest are used for the training, from run 1 to 100.

Stratified patient leave-out approach was selected to facilitate the model’s ability to more effectively identify the characteristics associated with the disease itself, rather than those specific to the patients. In a recent work on EEG disease diagnosis, Kunjan et al.^27^ highlighted that the cross-validation results without leave one subject out setup may provide an inaccurate representation, and hence misleading results. Experiments by corrupting the outcome variable both subject-wise and sample-wise pointed out that it is not reliable to use a cross-validation setup without leaving out subjects, as it grasps the patients’ characteristics. In another similar work on patients with congestive heart failure, Isler et al.^28^ evaluated the efficacy of cross validation methods on five distinct classifiers. Their findings revealed that the leave-one-out cross-validation method exhibited both the highest average performance and the lowest standard deviation values.

## Results

Of the 436 patients, 194 patients were female (44.4%). The most common condition was Tetralogy of Fallot (39.9%), followed by Atrial Septal Defect (17.6%) and Pulmonary Atresia with VSD (16.7%). Mean ECG age was 33 years (SD 11.7 and 75-25% IQR (40,23)). In several ECGs there were overlaps between individual lead signals on the original PDF documents, as can be seen in Figure 2. This included lead labels as well as large QRS complexes impinging on the lead displayed below it, in particularly the precordial leads (V2-V6). We developed an algorithm to successfully overcome this issue using the vector drawings on the original ECG PDF documents. Sample extraction results for the leads V4 and V5 can be seen in Figure 2.

Other artefacts were overcome without further user intervention, as shown in Figure 6 on another patient. We compared our work with prominent open-source approaches found in the literature to digitise ECGs like Paper-ECG^26^. Figure 6 demonstrates the advantages of our methods by comparing ECG segment extraction of the two algorithms side-by-side, respectively. Original signals from the corresponding ECG document are shown in Figure 6a, for leads v2 and v3. All the original signals have some difficulties like text overlaps of lead names and letter notes, signal overlaps between different leads. Those difficulties may lead the digitisation fail, as detailed in the following examples. While initial QRS complex in Figure 6a for v2 is distorted and abbreviated in a vertical direction in Figure 6b because of the lead name overlapping with the signal, our algorithm can correct this in Figure 6c. Figure 6d shows overlapping QRS complexes from the above lead impinging on the QRS complexes of the present lead. Our algorithm also corrects this in Figure 6e. Similarly, the baseline shift artefact in Figure 6d is corrected as shown in Figure 6e.

**Figure 6:**
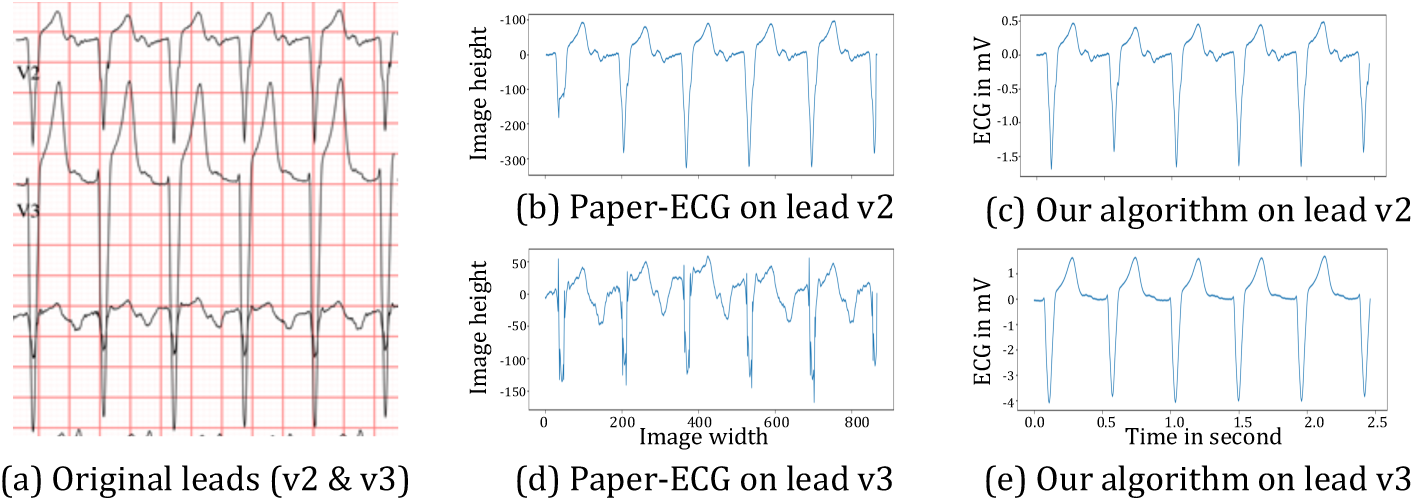
Comparison of our digitisation results of the vectorised graphic-based ECGs. On the left panel, original signals from the corresponding ECG document are shown for the leads v2 and v3. On the middle panel, digitisation results of Paper-ECG^26^ are displayed. On the right panel, digitisation results of our algorithm are displayed for the same leads of v2 and v3, respectively.

Bland-Altman plots are plotted to compare the measurement of the three variables using two different algorithms, vendor and our implementation. The mean of the two measurements is plotted on the x-coordinate while y-axis is the difference between the two algorithms, to show the agreement or disagreement between the two results. PR interval, QRS duration and ventricular rate are calculated and plotted in Figure 7a, 7b and 7c , respectively. For all the calculations, similar techniques as in the vendor manual^22^ were implemented, as closely as possible. Ventricular rates derived from the digitised ECG correlated well with the original vendor rendered ECG, as can be seen in Figure 7a. Similarly, in Figure 7b and 7c, the correlation between vendor calculated and digitised ECG calculated values for both QRS duration and PR interval is displayed respectively. Plots depict the agreement between the vendor calculated values on original signals and user calculated values on extracted signals. It also shows that there is no bias as the mean difference between two measurements is not consistently positive or negative. Pearson correlation coefficients are also reported along with the two-sided p-value, to show that there is strong correlation between the results. We also performed a null hypothesis test to calculate the significance of the correlation coefficient and to decide whether the relationship between the results is strong enough to be used to model the relationship. Null hypothesis assumes that the correlation coefficient is not significantly different from zero, and hence there is not a significant relationship between the variables. As the p-value is less than the chosen significance level (α = 0.05), we reject the null hypothesis. It indicates that there is sufficient evidence to conclude that there is a statistically significant correlation between the two results.

**Figure 7:**
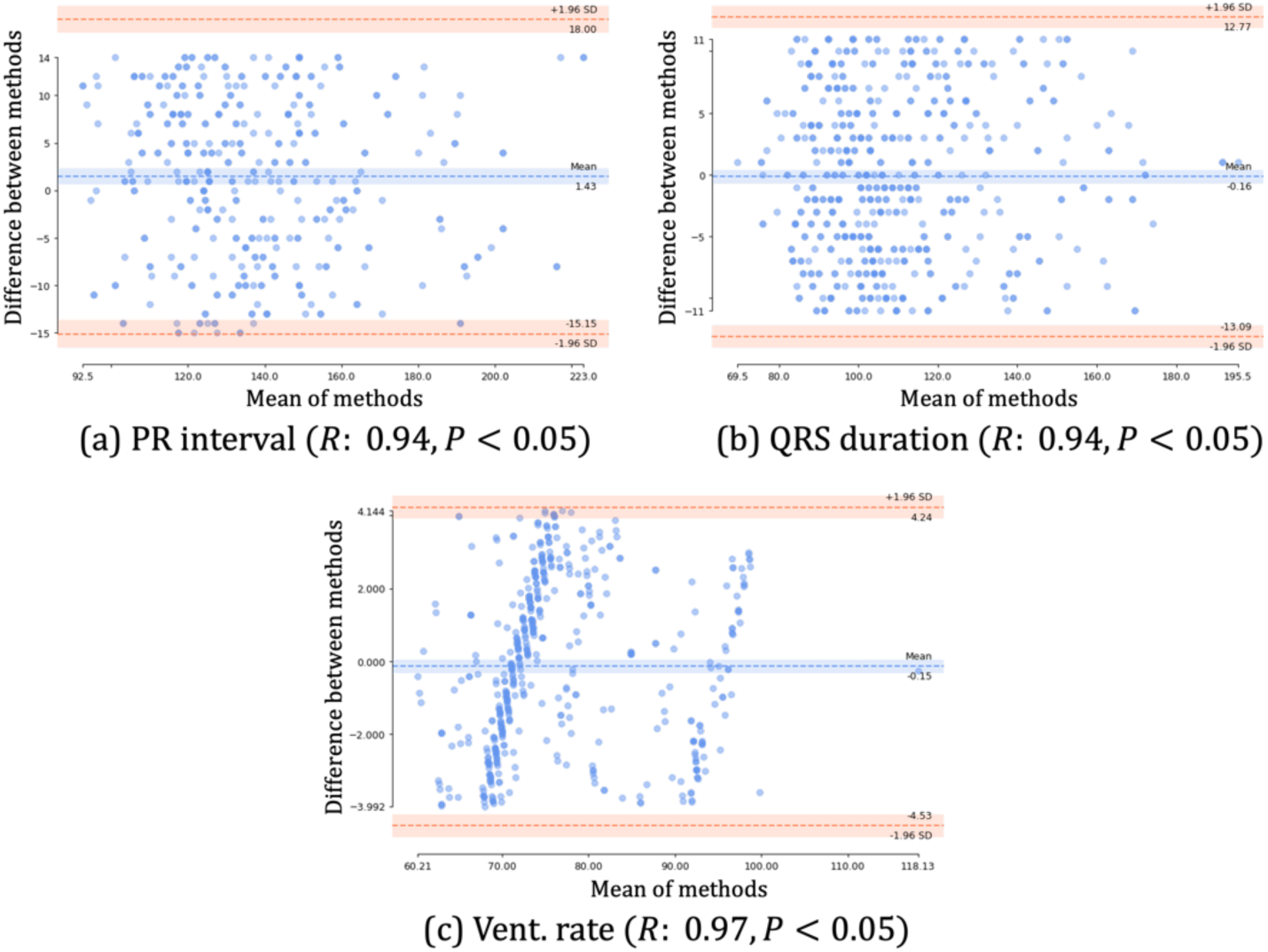
Bland-Altman plots between vendor and extracted values. PR interval, QRS duration and ventricular rate are calculated, and Bland-Altman plots are plotted in Figure 7a, 7b and 7c , respectively. Plots depict the agreement between the vendor calculated values on original signals and user calculated values on extracted signals. It also shows that there is no bias as the mean difference between two measurements is not consistently positive or negative.

Pre and post R-peak alignment differences are shown in Figure 3, along with the mean indicated by the black line. Improvements in consistency of machine learning performance were observed with the help of alignment on R-peaks, and the aligned data were much more successful in classification results.

The ability of the model to predict diagnosis accurately is displayed in Figure 8. Figure 8a, shows the performance of the classification on the AUC (Area Under the Curve) plot. It is a metric for assessing a binary classifier’s capacity to differentiate between classes by demonstrating the true positive against the false positive rate. As AUC scores are typically used in binary classification, One-vs-Rest (OvR) strategy is employed to evaluate AUC scores for each class separately. For example, the diagonal dashed line depicts the AUC curve of a random predictor which has a score of 0.5.

**Figure 8:**
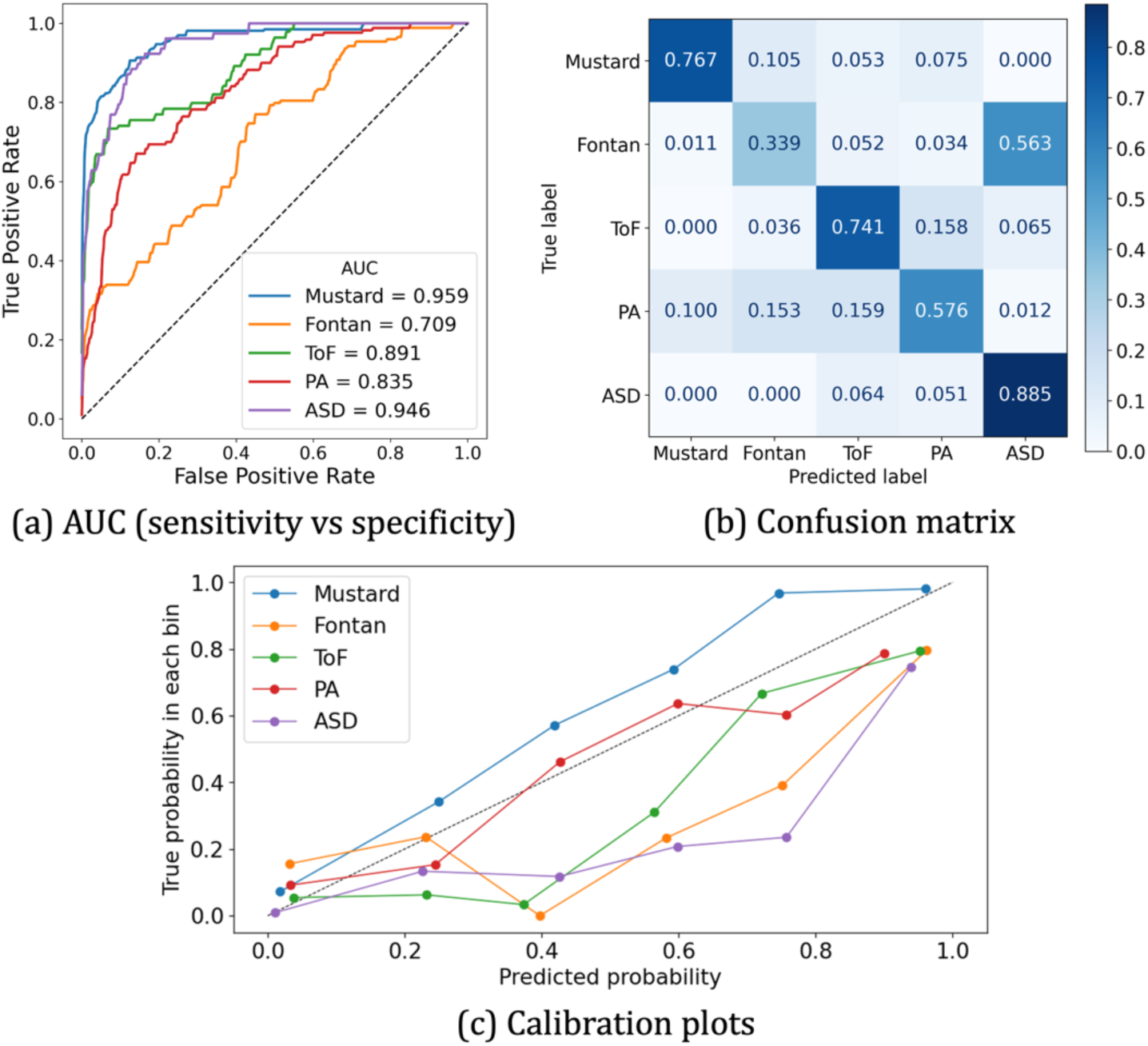
Classification performance of the SVM model. On the upper left panel, the average Area Under the Curve (AUC) scores of all 100 runs for each class are displayed separately. On the upper right panel, confusion matrix of all 100 runs combined is displayed to highlight model’s prediction vs true label. On the lower panel, calibration plots for each class are displayed separately. (**ASD:** Atrial Septal Defect, **PA:** Pulmonary Atresia, **TOF:** Tetralogy of Fallot).

Moreover, the confusion matrix for the SVM model can be seen in Figure 8b. Confusion matrix helps to understand the classes that are misclassified by presenting the model’s prediction summary in matrix form, showing how many predictions per class are correct and how many are incorrect. The model predicted a diagnosis of ASD accurately in 88% of cases, dTGA with a Mustard operation in 76%, ToF in 74%, PA in 57%, whereas in single ventricle circulations with a Fontan circulation only 34% of cases. All the classification metrics, including precision, sensitivity, specificity and F1 score, are reported in Table 1.

**Table 1:** Classification metrics for all the classes.

|  | Precision | Sensitivity | Specificity | F1-score |
| --- | --- | --- | --- | --- |
| **Mustard** | 0.915 | 0.966 | 0.767 | 0.834 |
| **Fontan** | 0.5 | 0.909 | 0.339 | 0.404 |
| **ToF** | 0.652 | 0.92 | 0.741 | 0.694 |
| **PA** | 0.653 | 0.92 | 0.576 | 0.612 |
| **ASD** | 0.388 | 0.854 | 0.885 | 0.539 |

## Discussion

Electrocardiograms (ECG) are powerful tools when used at a population scale to identify poor cardiovascular outcomes. ECGs can be used to provide a sense of the physiological and structural condition of the heart, while also providing valuable diagnostic clues. Currently analysis of large-scale data requires access to the original ECG datasets, which is embedded in a codified fashion within the vendor analysis software and is not readily accessible for analysis. Instead, clinicians tend to use manual evaluation of printed ECGs or review of ECG PDF documents for interpretation. Digitisation of such 12 lead ECGs as we have been able to demonstrate in this manuscript, is potentially very helpful in facilitating larger scale research including risk stratification, cross institutional research, and in registry data recording.

There are only a few studies on CHD (for adults or children) that focus on ECG analysis and classification using machine learning techniques due to the lack of diverse and large labelled data. Most previous works were implemented as a binary problem to detect whether a patient has the condition or not. For example, Du et al.^15^ proposed to use ECG for CHD screening in children. On the other hand, Khan et al.^16^ proposed to identify common congenital heart defects and distinguished them from normal ECGs. The authors proposed to extract ECG features such as mean, Route Mean Square (RMS), peak to peak and Signal to Noise Ratio (SNR) for prediction. Finally, Diller et al.^10^ proposed a deep learning architecture to categorise diagnostic group, disease complexity, and New York Heart Association (NYHA) class in a large cohort of patients with adult congenital heart disease only including some ECG parameters like resting heart rate, QRS duration, and QTc duration along with laboratory and exercise parameters.

In this work, we suggest linking ECG data with clinical letters and subsequently extracting accurately labelled digitised ECG waveforms for further analysis with machine learning algorithms. To our knowledge, this is the first time that ECG data from CHD patients are extracted from PDF documents and labelled automatically. Our proposed framework does not require additional user manipulation and we have extensively validated it by estimating ECG features, such as PR interval, QRS duration and ventricular rate and comparing the values with the vendor corresponding values.

There are several digitisation algorithms implemented by different researchers and vendors^19,20,26^, but to our knowledge all of them work on pixelated images captured from the ECG PDF documents, or from the scanned ECG papers. This usually results in quality loss, text along with the ECG waveforms overlap and the digitisation process fails to accurately depict the original signal. Here we used open-source tools to extract ECG vectorised graphical information from the PDF documents. In this way, we were able to reconstruct the signal accurately.

The digitised ECG overcomes artefacts present in the Paper-ECG^26^ digitised ECG traces in Figure 6, and accurately corrects these without any user intervention. For the first time, we demonstrate the ability for a relatively simple machine learning model to accurately predict diagnosis in the selected five conditions including Atrial Septal Defect, Mustard, Single Ventricle physiology with a Fontan circulation, Pulmonary Atresia and Tetralogy of Fallot. This preliminary research application and utility demonstrates encouraging potential for the technique. We believe that digitisation of the ECG will facilitate further research used in large datasets that can use PDF formats of the ECG.

The ability of the algorithm we developed to correct artefacts on the original ECG document was particularly gratifying. We demonstrate for the first time, the ability to correct for overlapping QRS complexes, baseline drift in the ECG, and text overlapping QRS or other parts of the ECG traces in patients with CHD. Though there are open-source works in the literature to digitise ECGs like Paper-ECG^26^, all such software currently needs user intervention to locate all the 12-leads with a corresponding bounding box. With our automated approach, we can not only digitise but also export the corrected ECG waveforms to PDF documents to avoid overlapping and allow clearer interpretation. So, it allows us to reconfigure the 12 ECG leads from the PDF raw data including individual lead vector data rather than having just an image for each lead as we are able to capture all the vectors on the PDF document.

We validated the ECG digitisation process by comparing vendor rendered ECG data like QRS duration, ventricular rate and PR interval between the original ECG and the digitised version. We were able to demonstrate a strong correlation between the measurements. Inherent variability in the intervals is expected as the techniques used by the vendor to measure particular intervals vary considerably. For example, some vendors use only simple band-pass filtering while others use template matching to detect QRS complexes.

In our case, raw ECG data was not readily available even for data analysis purposes. This is the main reason why we implemented such a digitisation pipeline, to enable further research on the raw data. We are also aware that this is not always the case, depending on the agreement with the provider. As they do not support for such research, not guarantee to access raw data for us. Our digitisation pipeline can be easily updated for different PDF structures/layouts of ECGs. We have tried several open-source works and non-commercial toolboxes to digitise ECGs (like Paper-ECG^26^), but the results were poor on the patients with CHD. ML model was a way to prove the algorithm works and it was sufficient for a smaller set of patients, more like a proof of concept.

We were able to use machine learning to train a relatively simple SVM model to recognise underlying cardiac anatomy such as in Tetralogy of Fallot, transposition of the great arteries with an atrial switch procedure, and patients with single ventricle physiology and a Fontan procedure. We demonstrated relatively poor discriminative ability of the model to successfully identify patients with a Fontan. Perhaps this is not unexpected given the enormous variety of underlying anatomic variation seen in patients with single ventricle physiology. This encompasses a wide range of anatomic diagnoses ranging from patients with a hypoplastic ventricle, to an almost non-existent right ventricle, and includes variations in ventricular dominance from left to right, and also position of the individual ventricles may vary enormously. We also had relatively few ECG data to analyse for individual lesions, and this could be greatly enhanced with larger scale datasets which is the next step in this research process.

### Limitations

The process currently employed to analyse ECG documents still requires manual deposition of the relevant 12-lead ECG documents into an analysis folder. Furthermore, despite the encouraging findings of this research, our total patient numbers were still relatively small, hindering more robust training of the ML algorithm. Automation of this process would aid efficiency. Finally, our proposed method has been developed on PDF documents that encode the ECG waveforms in a vectorised format. Therefore, our method cannot improve the extraction of ECG waveforms when ECGs had been digitised as pixelized images.

## Conclusions

We have adapted an extraction algorithm to digitise ECG PDF documents accurately without any user intervention. Our digitisation algorithm not only extracts ECG leads accurately but also provides further flexibility allowing the ECG PDF document to be reconstructed as requested. Also, using a rather simple machine learning algorithm, we demonstrated promising results on 12-lead ECG classification of anatomical diagnosis in congenital heart disease. Classification yields better results in terms of class distinction/discrimination on the aligned ECG signals, and it will be further tested on deep learning models to improve classification performance. Next steps would be to use this approach for risk stratification of mortality/cardiovascular hospitalizations and cardiovascular worsening in larger patient cohorts.

## Data Availability

All figures and code produced in the present study are available upon reasonable request to the authors.

---

**Central Illustration:**
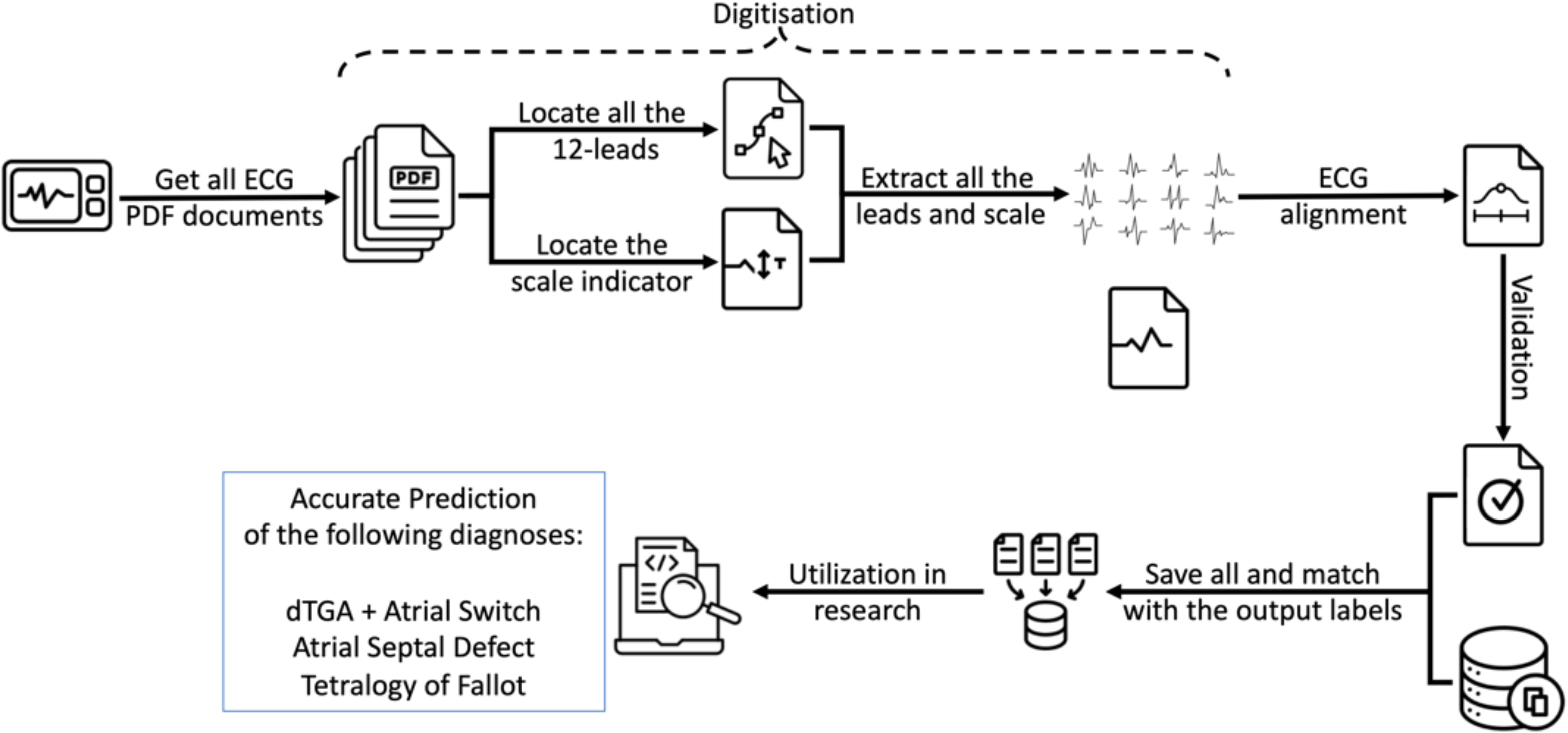
Overall workflow from digitisation to utilization in research Selected set of ECG PDF documents are placed in an analysis file. Then, digitisation is applied to extract all the leads and all the extracted data is stored for later use. Alignment and validation steps are also performed to ensure that there are no problems, and then all the data is matched with the corresponding output labels. After all these steps, it is now fully digitised and stored in a format that can be utilised in research.

